# Epidemiologically most successful SARS-CoV-2 variant: concurrent mutations in RNA-dependent RNA polymerase and spike protein

**DOI:** 10.1101/2020.08.23.20180281

**Authors:** Sten Ilmjärv, Fabien Abdul, Silvia Acosta-Gutiérrez, Carolina Estarellas, Ioannis Galdadas, Marina Casimir, Marco Alessandrini, Francesco Luigi Gervasio, Karl-Heinz Krause

**Affiliations:** Department of Pathology and Immunology, Faculty of Medicine, University of Geneva, Geneva, Switzerland; Department of Microbiology and Molecular Medicine, Faculty of Medicine, University of Geneva, Geneva, Switzerland; Department of Chemistry, University College London, London, UK; Institute for the Physics of Living Systems, University College London, London, UK; Institute of Structural and Molecular Biology, University College London, London, UK; Neurix SA, Geneva, Switzerland; School of Pharmaceutical Sciences, University of Geneva, Geneva, Switzerland; Institute of Pharmaceutical Sciences of Western Switzerland, University of Geneva, Geneva, Switzerland; Division of Infectious Disease and Dept. of Diagnostics, Geneva University Hospitals, Geneva, Switzerland

## Abstract

The D614G mutation of the Spike protein is thought to be relevant for SARS-CoV-2 infection. Here we report biological and epidemiological aspects of this mutation. Using pseudotyped lentivectors, we were able to confirm that the G614 variant of the Spike protein is markedly more infectious than the ancestral D614 variant. We demonstrate by molecular modelling that the replacement of aspartate by glycine in position 614 facilitates the transition towards an open state of the Spike protein. To understand whether the increased infectivity of the D614 variant explains its epidemiological success, we analysed the evolution of 27,086 high-quality SARS-CoV-2 genome sequences from GISAID. We observed striking coevolution of D614G with the P323L mutation in the viral polymerase. Importantly, exclusive presence of G614 or L323 did not become epidemiologically relevant. In contrast, the combination of the two mutations gave rise to a viral G/L variant that has all but replaced the initial D/P variant. There was no significant correlation between reported COVID mortality in different countries and the prevalence of the Wuhan versus G/L variant. However, when comparing the speed of emergence and the ultimate predominance in individual countries, the G/L variant displays major epidemiological supremacy. Our results suggest that the P323L mutation, located in the interface domain of the RNA-dependent RNA polymerase (RdRp), is a necessary alteration that led to the epidemiological success of the present variant of SARS-CoV-2.

## Introduction

The recent pandemic caused by the newly emerged SARS-CoV-2 coronavirus took the world by surprise. As opposed to previous coronavirus outbreaks such as SARS-CoV-1 and MERS, the new viral strain is not characterised by a particularly high case fatality rate, but rather by unprecedented infectivity and epidemiological success^1^. Indeed, the virus has so far largely resisted all attempts of eradication, despite stringent measures taken in many countries around the globe.

SARS-CoV-2 is an enveloped RNA virus, yet all coronaviruses differ from other RNA viruses in several respects. They have a large genome and a sophisticated life cycle which is still poorly understood. Also, coronaviruses have a proof-reading machinery, which confers a relatively stable genome, as compared to other types of RNA viruses^2-4^. Finally, given the magnitude of the pandemic, prevalent SARS-CoV-2 mutations leading to amino acid changes are relatively rare.

The SARS-CoV-2 transmembrane Spike glycoprotein (S-protein) is the most abundant protein of the viral envelope. It mediates attachment of the virus to the host cell surface receptors and a subsequent fusion between the viral and the host cell membranes to facilitate viral entry^5^. The S-protein is a trimeric class I fusion protein, which exists in a metastable pre-fusion conformation, divided into two main functional subunits responsible for binding to the host cell receptor (S1 subunit) and fusion of the viral and cellular membranes (S2 subunit, Figure 1a). The trimer exists mainly in two distinct conformational states resulting from an opening of the receptor binding domain (RBD) in the S1 subunit: ‘up’ or ‘down’ (Figure 1a). The ‘up’ conformation exposes the RBD to cell surface receptors of the host, which would otherwise be buried (‘down’ conformation) at the interface of each monomer^6^. This conformational change is required for proteolytic processing and/or fusion of the S2 domain with the host cell membrane. It is now understood that a variety of proteases, including the PC family of proteases, trypsin-like proteases and cathepsins are able to cleave the spike protein^7^.

**Figure 1.**
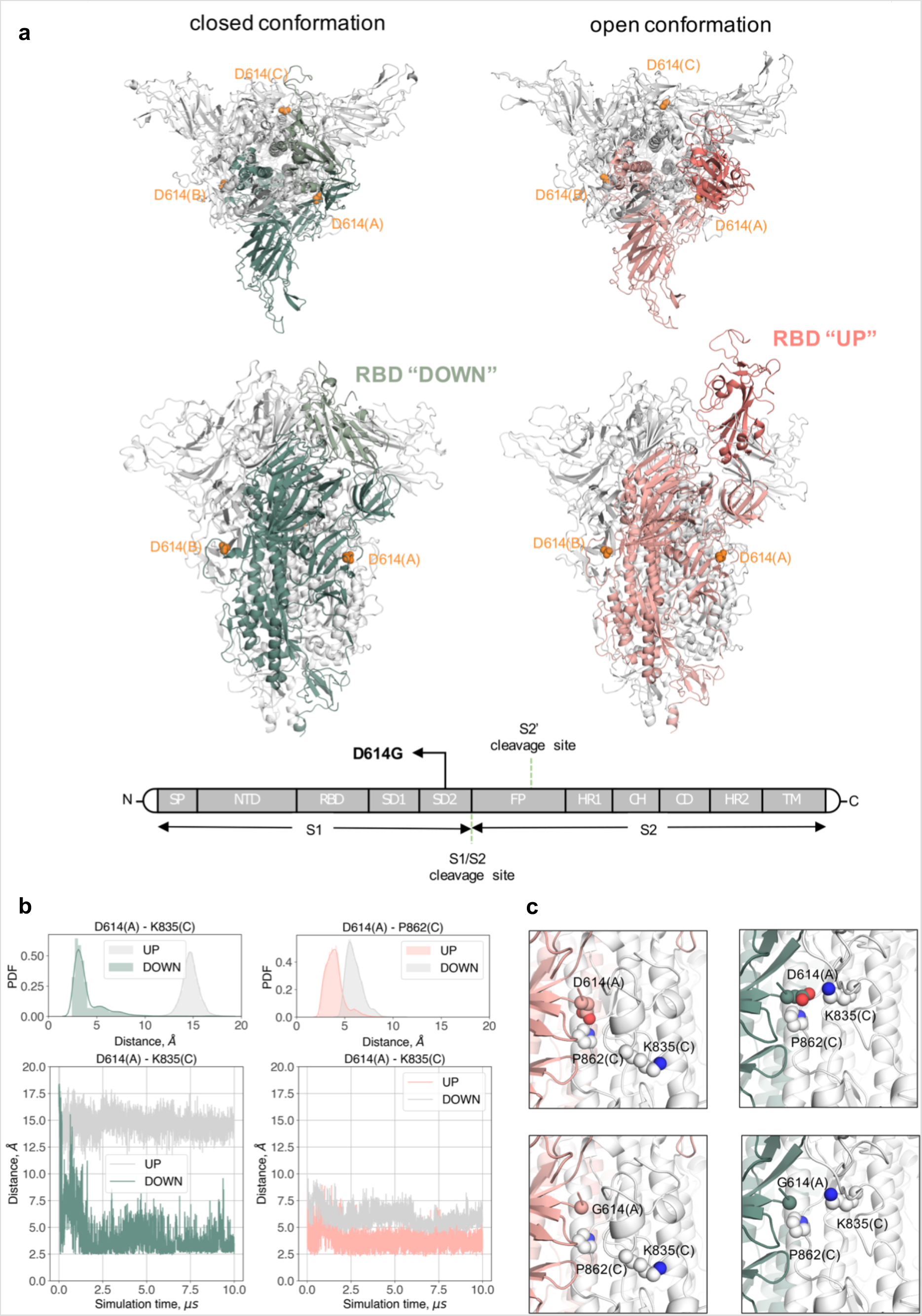
Structural implications of the D614G mutation in the S-protein. (**a**) Top and side views of the S-protein in the closed and open conformations. D614 in the three protomers is depicted in orange van der waals spheres. The RBD of protomer A in the *down* and *up* state is coloured in light green and dark pink, respectively. Graphical representation of the domain organisation of the S-protein: signal peptide (SP), receptor binding domain (RBD), N-terminal domain (NTD), subdomains 1 and 2 (SD1 and SD2), fusion peptide (FP), heptad repeat 1 (HR1), central helix (CH), connector domain (CD), heptad repeat 2 (HR2), transmembrane domain (TM). (**b**) Time series of D614 with K835 and P826 at the A-C interface over the course of the simulations initiated from the closed (green) and partially open (pink) conformations. The equivalent interactions of D614 at the B-C interface are given in grey. The probability distribution function (PDF) of the two interactions are displayed on the top of each timeseries. (**c**) Upper panels: interactions involving D614 for the closed (left) and open (right) conformation at the end of each simulation. The D614 residue of chain A is depicted in pink and green spheres and the interacting residues of chain C are represented in white spheres. Bottom-panels: interactions involving D614, when mutated to glycine, for the closed (left) and open (right) conformation at the end of each simulation. The Cα atom of G614 of chain A is depicted in pink and green spheres and the interacting residues of chain C are represented in white spheres.

The S-protein D614G mutation has received attention given that it could potentially alter the viral attachment, fusogenicity and/or immunogenicity. The mutation has been suggested to increase infectivity^8^ and/or the case fatality rate^9^. More recent studies have demonstrated that the G614 variant indeed enhances viral entry into cells^10,11^.

SARS-CoV-2 employs a multi-subunit replication/transcription machinery^12^. A set of nonstructural proteins (nsp) produced as cleavage products of the ORF1a and ORF1b viral polyproteins assemble to facilitate viral replication and transcription, making this machinery a potential target for therapeutic intervention^13^. RNA-dependent RNA polymerase (RdRp or nsp12) is a key player in the synthesis of viral RNA. The recently resolved crystal structure of SARS-CoV-2 nsp12 in complex with its nsp7 and nsp8 co-factors underlines the central role these non-structural proteins have in the replication and transcription of the virus^13,14^. The P323L mutation of the RdRp has received less attention; however, it has been suggested that this mutation might alter viral proof reading and thereby lead to an increased down-stream mutation rate^15^.

In this study, we investigated the emergence of a SARS-CoV-2 variant with concomitant mutations in the S-protein and RdR-polymerase. Both mutations are in strategically relevant sites of the respective proteins and hence candidates to alter the biology of the virus. We show that the G614/L323 viral variant, but not the individual G614 or L323 mutants, is epidemiologically highly successful and over the last months has largely replaced the original D614/P323 variant.

## Results

### S-protein D614G increases cell transduction of pseudotyped lentivectors

Cell entry of SARS-CoV-2 depends on binding of viral S-proteins to cellular receptors and subsequent priming by proteases of the host cell^16^. Our first aim was to establish cell lines that consistently express the angiotensin-converting enzyme-2 (ACE2) receptor, which is recognised by SARS-CoV-2; and relevant proteases, such as transmembrane serine protease 2 (TMRSS2), which cleaves the S-protein. As seen in Figure 2a and b, neither HeLa cells nor A549 cells expressed relevant amounts of ACE2 mRNA. Similarly, mRNA levels of TMRSS2 were almost undetectable. However, both cell lines express low to intermediate gene levels of furin and moderately high levels of cathepsin L (Figure 2). Furin is able to cleave the spike protein of SARS-CoV-2, while cathepsin L is able to cleave S-proteins from both SARS-CoV-1 and SARS-CoV-2^7^. In order to establish cell lines permissive for S-protein pseudotyped lentivectors, we transduced both HeLa and A549 cells with either ACE2 alone, TMPRSS2 alone or in combination.

**Figure 2:**
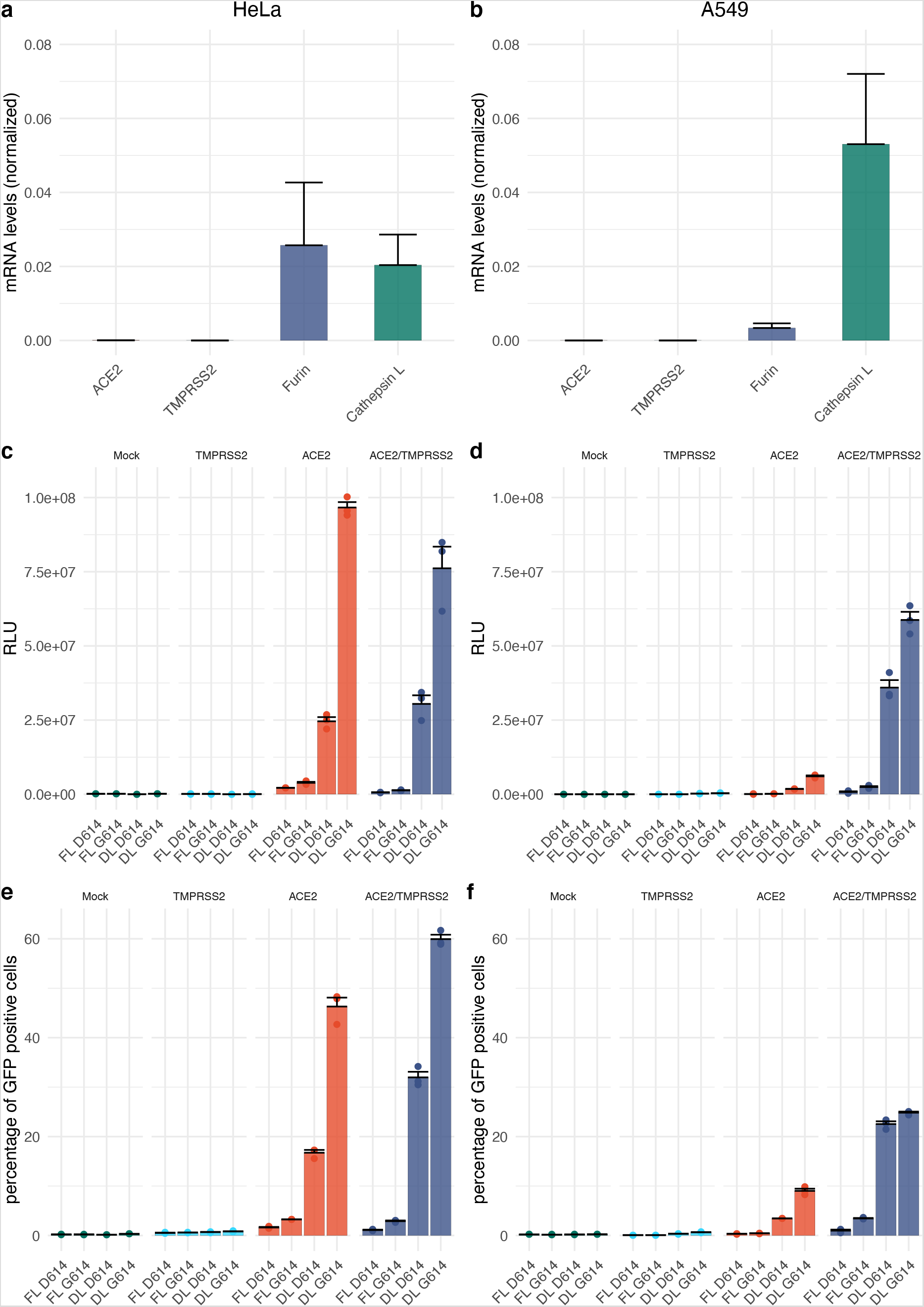
SARS-CoV-2 S-protein D614G variant shows increased infectivity. (a-b) Real-time RT-PCR for ACE2, TMPRSS2, furin and cathepsin L mRNA expression profiles in HeLa **(a)** and A549 **(b)** cells. **(c-f)** Spike pseudotyped lentiviral infectivity assays. Pseudotyped lentiviruses expressing Luciferase and GFP and pseudotyped with SARS-CoV-2-D614 or G614 full length or truncated version were used to transduce HeLa and A549 overexpressing or not either ACE2 and/or TMPRSS2. **(c-d**) Luciferase assay of HeLa **(c)** and A549 **(d)** at 5 days post transduction with 80 uL of SARS-CoV-2 spike pseudotyped lentivirus. (**e-f**) Percent of GFP+ assay of HeLa **(e)** and A549 (**f**) cells at 5 days post transduction with 80 uL of SARS-CoV-2 S-protein pseudotyped lentivirus analysed by Flow cytometry. The experiments were done in triplicates and repeated three times with three independent lentiviral stocks that was resulting from independent transfection. One representative is shown with error bars indicating SEM of replicates.

To understand the impact of the S-protein D614G mutation on infectivity, we produced S-protein pseudotyped lentivectors. Specifically, we used full length and C-terminally truncated D614 protein, as well as full length and C-terminally truncated G614 S-protein. We included the C-terminal truncation, as it has been shown previously to increase transduction with spike pseudotyped subviral particles, probably due to the removal of a retention sequence^17^. Our pseudotyped lentivectors expressed both a secreted luciferase, as well as GFP; therefore, measurements were performed by luminescence detection and flow cytometry.

As expected, exposure of wild type (mock-transduced) HeLa or A549 cells to the lentivectors described above did not lead to GFP or luciferase expression, which suggests that these cells were not permissive to transduction by our pseudotyped lentivectors (Figure 2c-f). Similarly, overexpression of TMPRSS2 by itself did not lead to detectable transduction. Using the D614 pseudotyped lentivector, low-level transduction of ACE2 and ACE2/TMPRSS2-expressing HeLa cells were observed, but which was hardly detectable with A549 cells. This changed markedly when the G614 pseudotyped vector was used, with there being significant transduction of both ACE2-expressing cell lines. Once again, transduction of ACE2-expressing HeLa cells was significantly higher than for ACE2-expressing A549 cells. Interestingly, overexpression of TMRPSS2 in combination with ACE2 expression led to an additive enhancement of transduction in A549 cells, which was not the case for HeLa cells. We hypothesized that our HeLa cell line most likely had high-level expression of a yet to be defined S-protein cleaving protease, and that overexpression of TMRPSS2 is not required for efficient transduction. In contrast, in A549 cells, proteolytic cleavage appears to be rate-limiting, and overexpression of TMRPSS2 is required to enhance transduction.

### D614G mutation alters the interactions around the point of the mutation

Recently, several trajectories from long molecular dynamics (MD) simulations of the S-protein of the SARS-CoV-2 virus have been released to the public domain (CC BY 4.0)^18^. These long simulations (10 s) were obtained in specialised supercomputers^19^. For the present analysis, we used two trajectories of the trimeric SARS-CoV-2 spike glycoprotein^18^. The first simulation was initiated from the closed, “down” conformation (PDB entry 6VXX), and the second one from a partially open, “up”, conformation (PDB entry 6VYB)^20^. The simulated structures contain modelled missing loops from the crystal structures and glycans^21^. Although no conformational transitions between the two states are observed during these long trajectories, the analysis of the dynamics of the key interactions around the point of mutation helps to better understand the effect of the latter.

The pairwise (all-to-all) root-mean-square deviation (RMSD) matrix of the S-protein shows for both closed and open simulations that the starting crystal structure relaxes quickly, leading to a different stable conformation with respect to the initial one due to the effect of the glycan dynamics (Supplementary Figure 1). The overall dynamics of the domains of the protein is different. While the S2 subunit and the N-terminal domain (NTD) of the S1 subunit are very stable for all three protomers in both the closed and open conformations, the RBD domain of protomer A exhibits a noticeable change between the two conformations. Specifically, its RMSD in the up conformation is higher than in the down one due to its solvent exposure (Supplementary Figure 2). It is worth noting that the RBD domain in the up conformation explores two states during the dynamics, as the presence of the glycans affects the initial crystal structure conformation. In the MD simulation with all three RBD domains in the *down* state (closed conformation), the D614 from chain A forms a salt bridge with K835 in the adjacent chain C (Figure 1b upper panel, green). In the open conformation, where the RBD domain of protomer A is in the *up* state, this salt bridge is lost and D614 in protomer A turns inwards facing the core of the S2 subunit forming hydrogen bond with P862 of protomer C (Figure 1b upper panel, pink).

The interactions involving D614, once the structures are fully relaxed after MD simulations, clearly reveal a different behaviour between the closed and open conformations. The D614G mutation will disrupt the formation of the D614(A)-K835(C) salt bridge in the closed conformation, but very little will change in the open conformation (Figure 1c). The D614G mutation stabilizes the open conformation of the S-protein as the pairwise RMSD correlation matrix is very stable (Supplementary Figure 2) even for the RDB (A) domain exposed to the solvent. A recent pre-print points in the same direction where negative stain electron microscopy reconstructions of the D614G form of the S-protein shows that the mutation favours the ‘up’ conformation, 82% population compared to 42% in the wild type^22^.

### Two abundant amino acid changes in SARS-CoV-2: S protein D614G and RNA-dependent RNA polymerase (RdRp) P323L

27,084 genomic sequences from GISAID (http://www.gisaid.org/)^23^, representing 87 countries over the time period of 24 December 2019 to 19 June 2020, were used to perform the epidemiological study. Using the original D614/P323 variant (NC_045512.2) as a reference, we identified two highly abundant SARS-CoV-2 mutations leading to amino acid changes: the D614G mutation in the S-protein, and the P323L mutation in the RNA-dependent RNA polymerase (RdRp). Figure 3a-b shows the weekly frequency of these mutations in the viral sequences over time (log scale), while Figure 3c-d shows the percentage of the respective mutants within a given week. Until mid- January 2020, both mutations were virtually absent in the available sequences. Thereafter, we can observe a notable increase in frequency of this mutant, with a virtually complete replacement of the original sequences by May 2020. A striking similar increase in frequency of both the D614G and P323L mutations should be noted.

**Figure 3.**
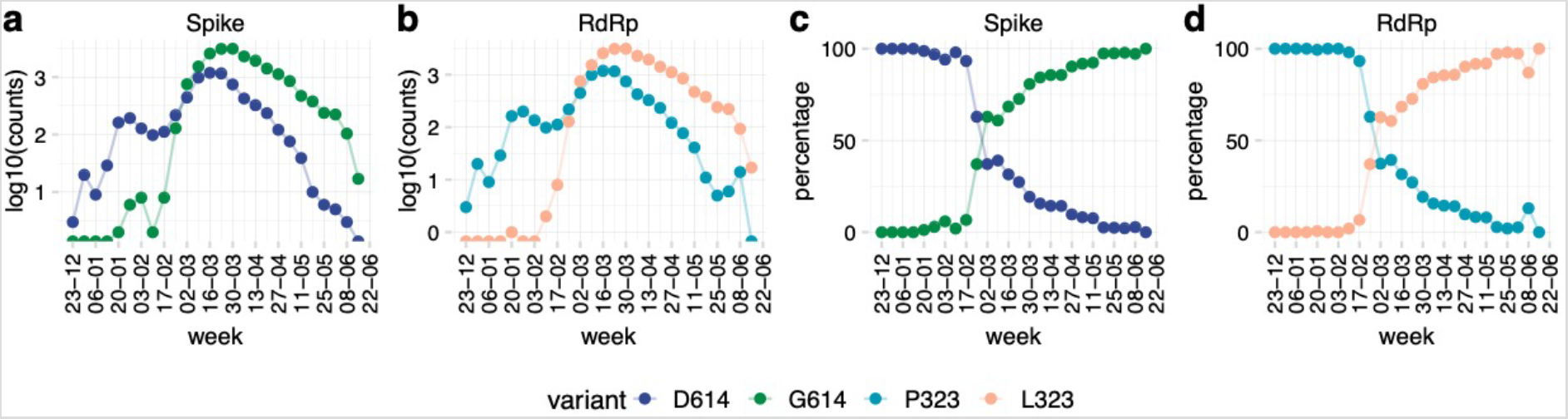
Two major protein variants of SARS-CoV-2: S protein D614G and RNA-dependent polymerase P323L. Available high-quality sequences of SARS-CoV-2 (27 084) were analysed for the D614 and G614 variants of the S-protein and the P323 and L323 of the RdR polymerase. Data are shown as total number of sequences per week (**a**, note the logarithmic scale) or percent of sequences per week (**b**).

### Co-emergence and concurrence of the D614G and the P323L mutations

Similar kinetics in the appearance of D614G and P323L led us to further analyse the relationship between the two mutations. Figure 4a shows the occurrence of the four possible variants: D614/P323 (i.e., the original D614/P323 variant), G614/P323 (mutation only in the S-protein), D614/L323 (mutation only in RdRp), and G614/L323 (mutations in both the S-protein and RdRp). In our dataset, these variants were first reported as follows: D614/P323 on 2019–12–24; G614/P323 on 2020–01–24, D614/L323 on 2020–03–10, and G614/L323 on 2020–01–24. Interestingly, only the double mutation G614/L323 was epidemiologically successful and presently accounts for ∼70% of all available SARS-CoV-2 sequences; while the original Wuhan sequence, accounts for ∼30%. The single mutants account for only 0.3% (G614/P323) and 0.1% (D614/L323) of all sequences. To investigate the emergence and spread of the strains, we analysed the weekly numbers of sequences (Figure 4b) and the cumulative number of sequences (Figure 4c) of D614/P323 and G614/L323 variants. There was an evident first wave from December 2019 to February 2020, which was strongly dominated by the D614/P323 variant. From March 2020, a second wave with a predominance of the G614/L323 variant can be observed. We also observed major differences between the variants in different countries (Figure 4d). For example, while D614/P323 was the largely dominant variant in Hong Kong, South Korea and China; G614/L323 was almost exclusively observed in countries such as Switzerland, Czechia and Argentina. We also note that the occurrence of single mutants (G614/P323 and D614/L323) continued to be rare.

**Figure 4.**
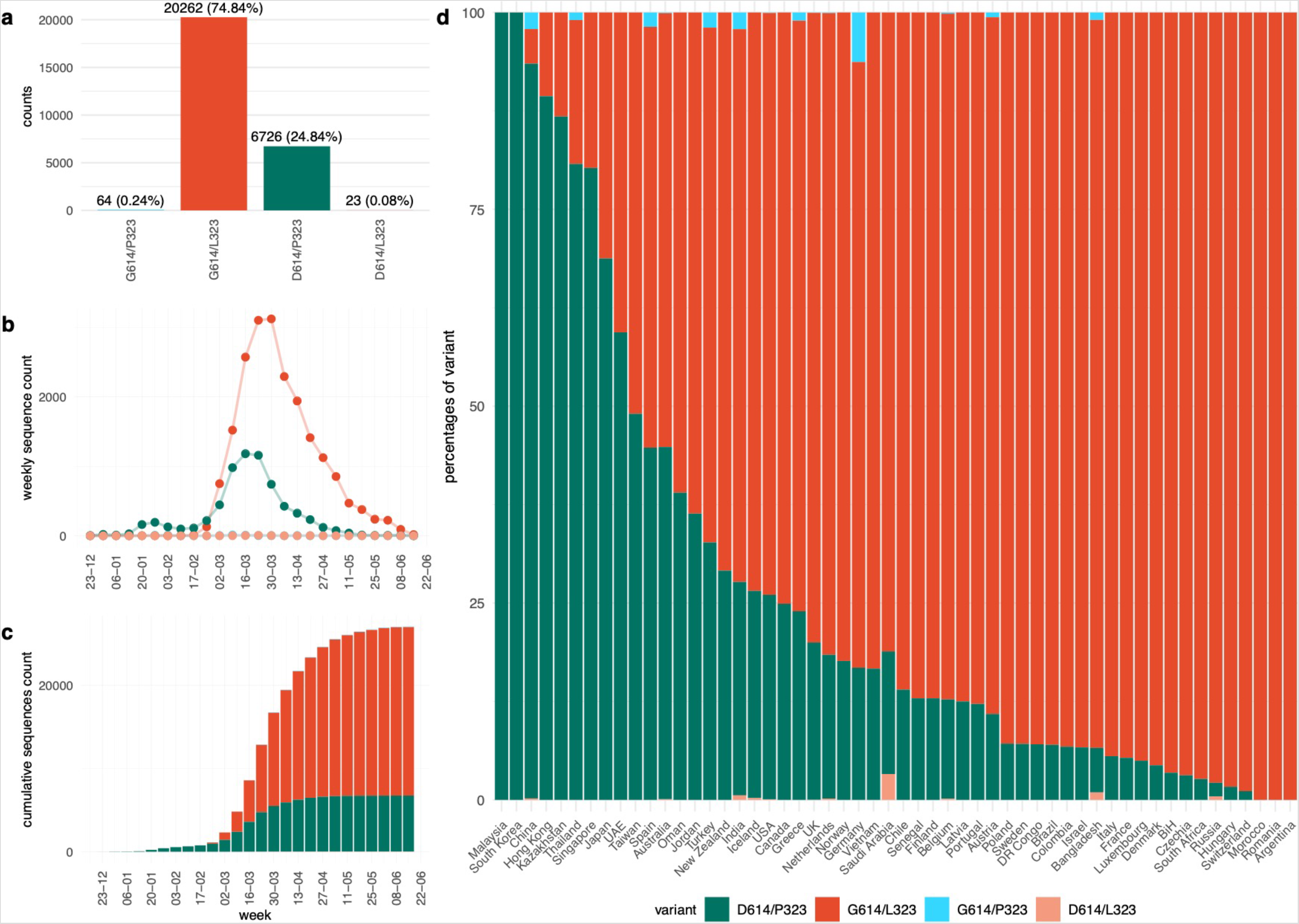
Temporal and spatial distribution of the four possible D614G and P323L combinations. Available high-quality sequences of SARS-CoV-2 (27 075) were analysed. **(a)** G614/L323 (double mutant) represented more than two-thirds, and D614/P323 (the original Wuhan sequence) about one-third of sequences; the single mutant variants G614/P323 and D614/L323 were very rare; **(b)** weekly number of variants over time; **(c)** cumulative number of variants over time. **(d)** percentage of variants in different countries; only countries with at least 20 sequences are represented.

To better understand the relationship between the dynamics and geographic aspects of the pandemic, we analysed the weekly and cumulative numbers of sequences (Figure 5) in different countries (only countries with at least 20 sequences were included in this analysis). At first glance, the D614/P323 variant can be observed as the dominant strain in the initial wave in Asian countries, while the G614/L323 variant was the main driver of the pandemic in other continents. However, in several Asian countries, including Taiwan, Thailand, Japan; the emergence of G614/L323 can be observed at later time points. Also, in many non-Asian countries, the pandemic started with the D614/P323 variant, followed by a strong predominance of the G614/L323 variant (e.g. Italy, UK, USA). Thus, country-by-country data do not suggest a simple geographic predominance or a bottleneck in the transmission that led to a random emergence of G614/L323 in non-Asian countries. Rather, it appears that G614/L323 is epidemiologically more successful than the original D614/P323 variant.

**Figure 5.**
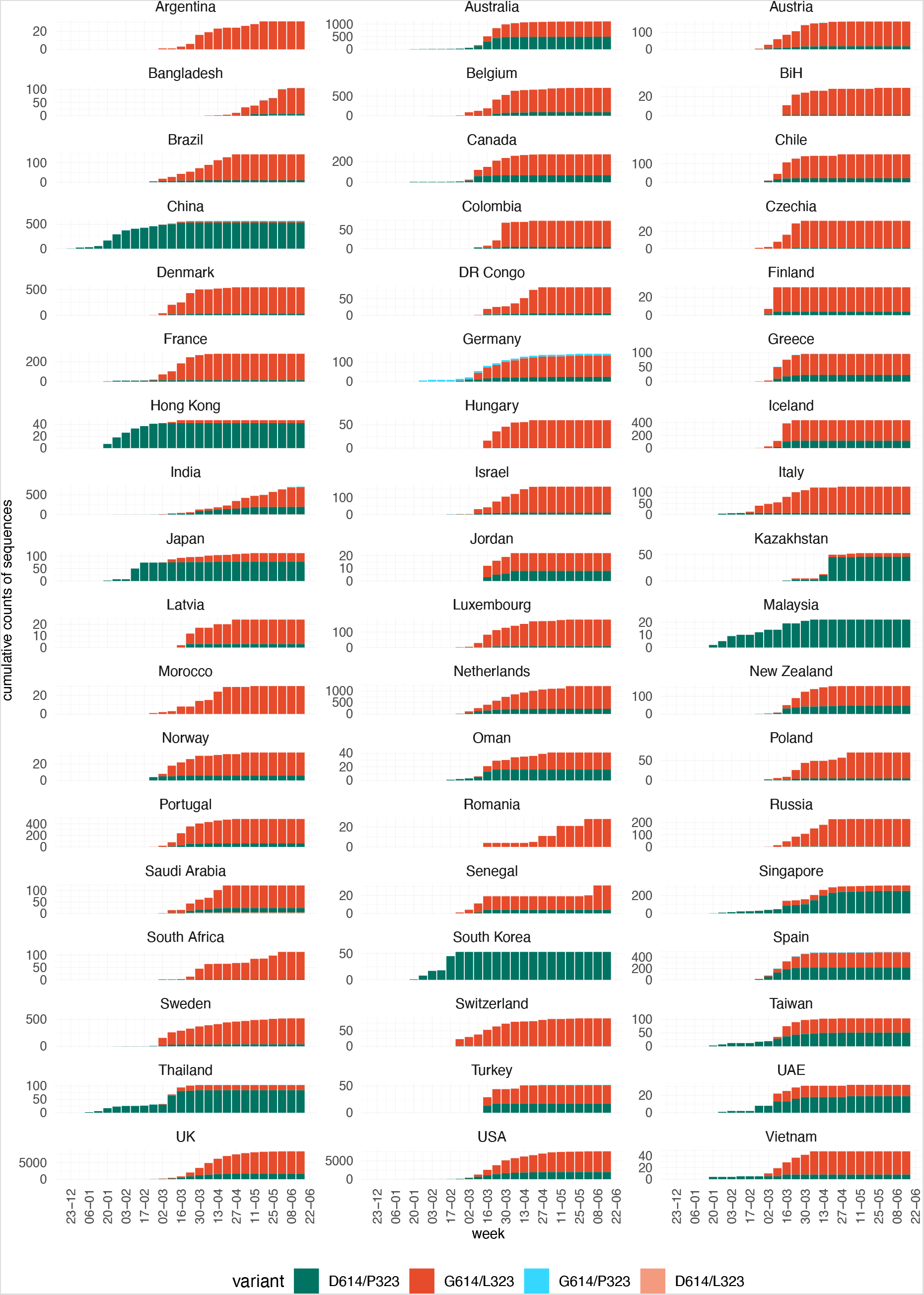
Temporal evolution of the four possible D614G and P323L variants in different countries. Available high-quality sequences of SARS-CoV-2 (27 075) were analysed for the different possible combinations of the D614 and G614 variants of the spike protein and the P323 and L323 of the RdR polymerase based on country of origin of the sequences. Only countries with at least 20 sequences were included in this analysis. Results are shown as cumulative number of sequences over time (the weekly number of sequences is provided as a Supplementary Figure 5).

We also investigated the case fatality rate in different countries as a function of the percentage of D614/P323 vs. G614/L323. We found no significant correlation between strain predominance and either deaths by million population (Figure 6a) or case fatality rate (Figure 6b). However, we cannot entirely exclude such a correlation, as there was a weak trend towards a higher case fatality rate in countries with a high percentage of the G614/L323 variant. For 1,845 sequences in our analysis, patient status was available from GISAID (http://www.gisaid.org/)^23^. These data are submitted in a text-free form (e.g. asymptomatic, home, hospitalised, released, recovered, live, deceased), but which can contain errors, redundant status descriptions and country specific biases. Also, compared to the total number of sequences, only a few genomes had a status attribution. Bearing in mind these limitations, the data can be used to establish a first approximation between viral genotype and patient status. Patients with the double mutation variant were more often attributed a status of “deceased” and “hospitalised”; while patients with the original D614/P323 variant, were more often attributed a status of “asymptomatic” (Supplementary Figure 3).

**Figure 6.**
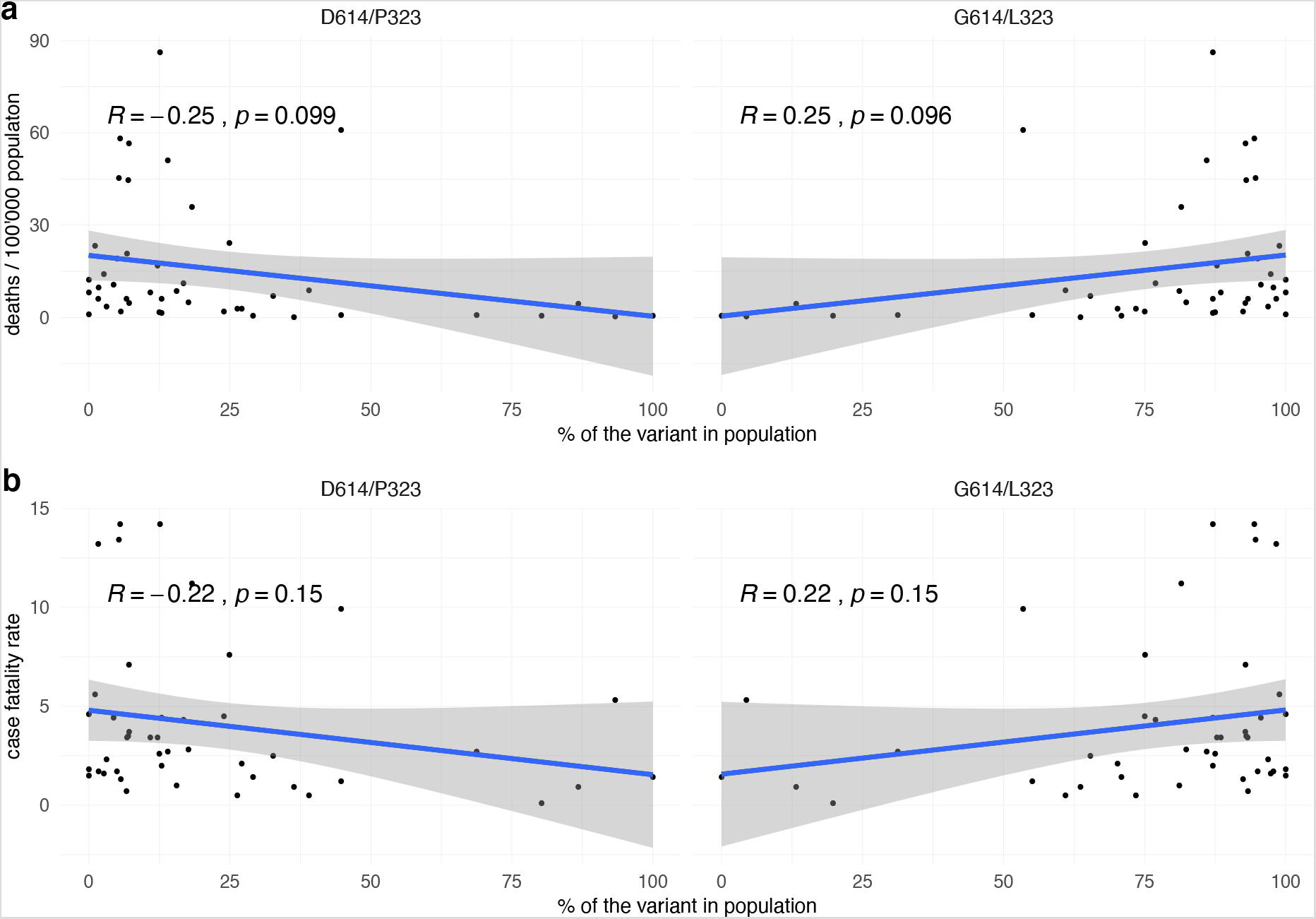
SARS-CoV-2 variants: mortality, case fatality rate, and patient status. Relationship between the percentage of the two main SARS-CoV-2 variants (D614/P323 and G614/L323) in different countries and **(a)** number of deaths per 100 000 population and **(b)** case fatality rate in the respective countries. Numbers of deaths per 100 000 population and case fatality rates were taken from the Johns Hopkins Coronavirus Resource Center (http://coronavirus.jhu.edu/data/mortality) at 4^th^ of august 2020.

We next investigated the epidemiological parameters in more detail. In 66 countries, sequences of both the D614/P323 variant and the G614/L323 were reported. In 37 countries, the D614/P323 variant was reported before the G614/L323 (Figure 7a, light blue bars); while in 24 countries, the double mutation was reported first (Figure 7a, dark blue bars). In five countries, both variants were reported for the first time on the same date. Note that even in countries where the D614/P323 variant was reported more than 20 days before the G614/L323, ultimately a clear predominance of the G614/L323 variant was observed (e.g. Belgium, USA, France). We therefore decided to compare the spread of the two dominant strains and the single mutant variants in different countries with respect to the total number of sequences collected within 14 days following the emergence of each respective strain (only countries with at least five sequences at day 14 were included). As shown in Figure 7b, the emergence of the single mutant strain was slow and of low efficiency. The D614/P323 variant shows a relatively slow emergence, which increases mostly after seven days. In contrast, the G614/L323 strain emerged very rapidly and efficiently. When comparing the rate at which the strains emerge within the given countries, the difference between the D614/P323 variant and the G614/L323 strain was statistically significant (p-value < 2.2e^−16^; Wilcoxon signed rank test with continuity correction).

**Figure 7:**
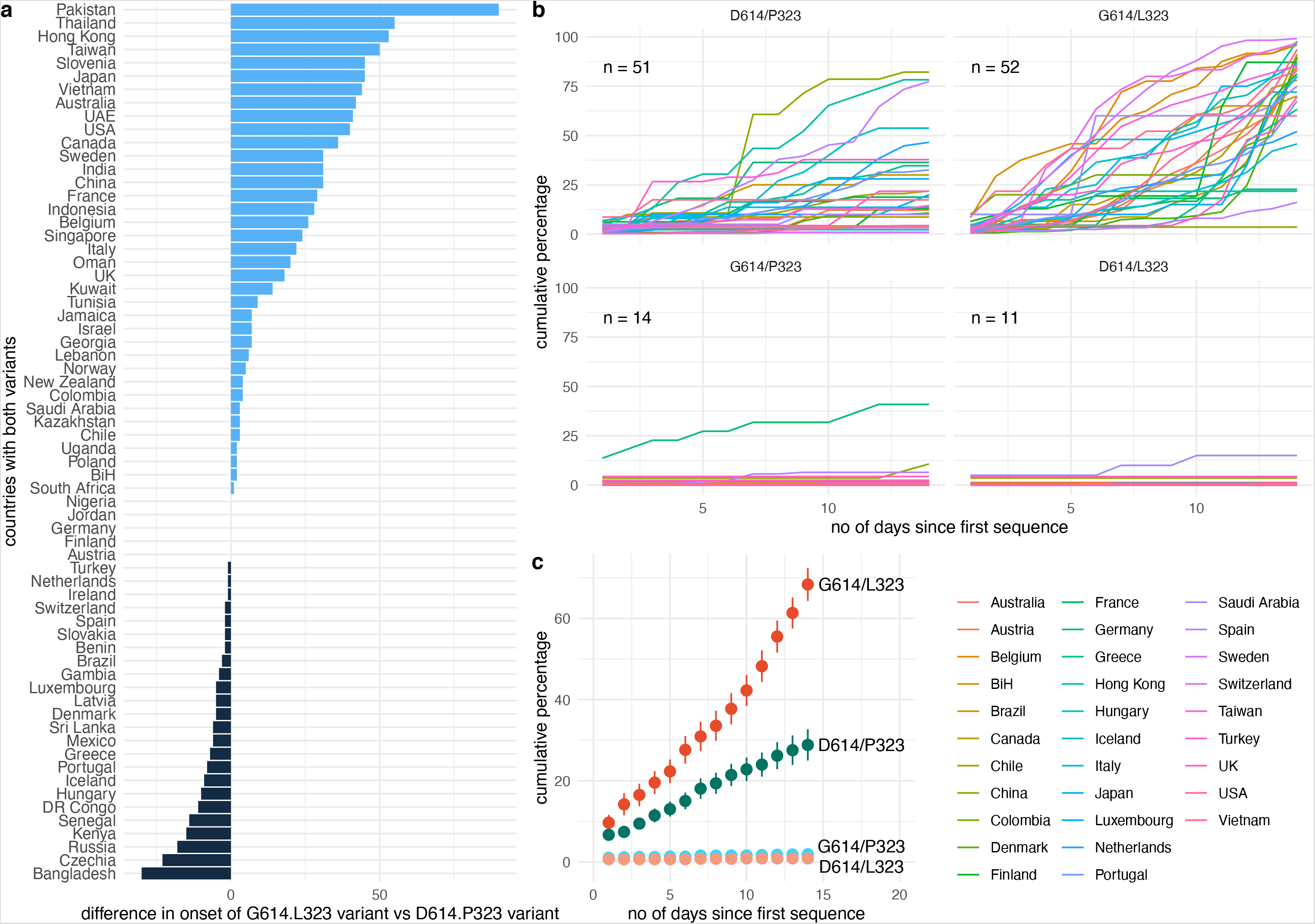
Comparative expansion of SARS-CoV-2 variants in different countries: (a) time of first reported D614/P323 sequence versus first reported G614/L323 sequence (blue bars: D614/P323 appeared before G614/L323; black bars: D614/P323 appeared before G614/L323). **(b)** percent of sequences of a given variant in the 14-day time period after its first appearance in a given country (100% is total number of sequences during the 14-day time period). Only countries with at least 5 sequences collected during 14 days after the appearance of first sequence are used (**c**) mean +/− SEM of emergence of the variants in different countries with at least 5 sequences within the first 14 days.

### P323L mutation of the SARS-CoV-2 RdRp polymerase

The SARS-CoV-2 polymerase RdRp has an overall architecture that resembles the one seen in SARS-CoV-1 and consists of a nsp12-nsp7-nsp8 complex. The structure of the SARS-CoV-2 nsp12 contains a cupped, right-handed RdRp domain linked to a nidovirus RdRp-associated nucleotidyl-transferase domain (NiRAN) via an interface domain. The RdRp domain adopts the conserved architecture of the viral polymerase family 4 and is composed of three domains: a fingers domain, a palm domain and a thumb domain (Figure 8). The nsp12 is stabilised by a nsp7-nsp8 heterodimer, which is packed against the thumb-finger interface, and a single nsp8 monomer interacting with the finger and interface domains. The P323L mutation is located on the interface domain of nsp12, and specifically on the loop that connects the three helices of the interface domain to the three β-strands of the same domain.

**Figure 8:**
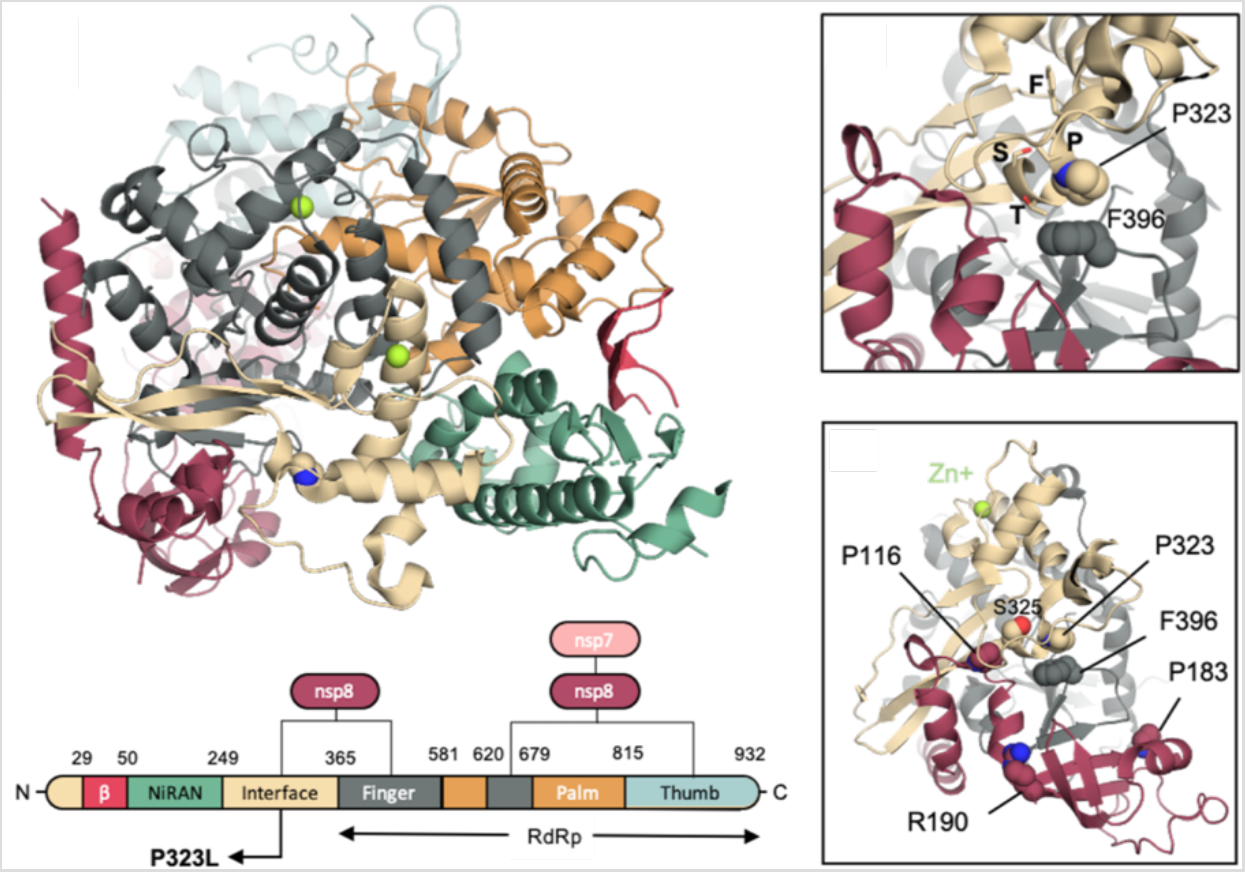
Structural implications of the D614G mutation in S-protein. (a) Domain organisation of SARS-CoV-2 nsp12 in complex with nsp7 and nsp8 cofactors. The P323 is depicted in beige spheres. **(b)** Close up around P323. The residues of the ^321^FP**P**TS^325^ stretch are depicted in sticks, while F396 that P323 interacts with is depicted in spheres. **(c)** Residues P116, P183 and R190 of the nsp8 that has been reported to be crucial for the nsp8/nsp12 interaction in SAR-CoV-1 are depicted in red spheres.

Although P323 engages only in a single interdomain interaction with F396 of the finger domain (Figure 8), L323 is expected to affect the local dynamics of the regions around the point of the mutation. The conformational freedom of the proline rich ^321^FPPTS^325^ stretch, where the P323L mutation lies, is expected to be limited due to the pyrrolidine ring of the two Pro residues, which provide a degree of local structural rigidity. Phe has been shown to be over-represented in sequence positions immediately following Pro-Pro motifs in protein turns and NMR data have shown a strong Pro_i_-Phe_i+2_ C_α_Η – π interaction that restricts the rotation of the Phe side chain^24^. Therefore, although L323 will still in principle be able to interact with F396, the mutation is expected to increase the flexibility of this region.

A mutagenesis study has confirmed the functional role of nsp7 and nsp8 in SARS-CoV-1 replication^25^. Specifically, Subissi et al. identified three nsp8 residues (P116, P183, and R190) that are critical for the nsp8/nsp12 interaction *in vitro* and *in vivo*, and the reported impaired SAR-CoV-1 polymerase activity in mutant variants underlines the key role of these residues in the nsp8/nsp12 association and subsequent SARS-CoV-1 RNA synthesis^25^. Although similar experiments have to be carried out for the case of SARS-CoV-2, R190 and P116 are very close in space to the point of the P323L mutation, and P116 interacts directly with S325 of the ^321^FPPTS^325^ stretch (Figure 8). Therefore, we anticipate the P323L mutation to have an impact on the nsp8 association. The level of association of nsp8 has been shown to be an important factor for obtaining a high RNA polymerase activity in SARS-CoV-1^25^, and therefore enhanced interaction between the nsp12 and nsp8 upon the P323L mutation may give an evolutionary advantage to the virus.

## Discussion

Compared to DNA-based organisms, RNA viruses are generally believed to replicate their genetic material with lower accuracy, as they lack the necessary proofreading and correction mechanisms^26^. Given the high mutation rate seen in viruses, the mutations that give an advantage to the virus and, therefore, are successful, are fine-tuned through evolutionary pressure. Here, we have analysed the structural and functional impact of the widely discussed S-protein mutation D614G and investigated whether it provides by itself an epidemiological advantage to the SARS-CoV-2. We demonstrate that indeed, this single amino acid substitution from aspartate to glycine at position 614 significantly enhances the infectivity of pseudotyped lentivectors. Through molecular modelling, we suggest that the G614 amino acid substitution destabilizes the closed conformation, while it does not alter the stability of the open one. The transition towards the open state is thought to be required for viral fusion,^5^ and thus a structurally favoured open state is in line with the higher infectivity demonstrated by our biochemical assays.

However, epidemiological data suggests that the D614G mutation is not exclusively responsible as a driver mutation of SARS-CoV-2. The time series for emergence of the D614G and P323L mutations indicates that the S-protein mutation (D614G) evolved simultaneously with P323L in RdRp. We observed small clusters of the G614/P323 single mutant variant on two occasions: February 2020 in Germany, and June 2020 in India. Yet, these clusters were always transient and failed to make major epidemiological contributions. Thus, our data suggest that the P323L mutation of the polymerase plays an important role in the present epidemiological success of D614G and SARS-CoV-2 in general.

Although more experiments are needed to understand the underlying biological relevance of the two mutations, we speculate that the P323L mutation attenuates the clinical presentation of the SARS-CoV-2. Firstly, based on its location, we expect the P323L mutation to negatively impact the nsp8 association. The level of association of nsp8 has been shown to be an important factor for obtaining high RNA polymerase activity in SARS-CoV-1^25^, and thus P323L is likely to attenuate polymerase activity in SARS-CoV-2. Secondly, experience from SARSCoV-1 and MERS suggests that a more aggressive virus is – from a public health perspective – easier to eliminate (it is easier to diagnose acute disease with few asymptomatic carriers). Indeed, SARS-CoV-1 and MERS resulted in higher case fatality rates relative to SARS-CoV-2, but were both epidemiologically less successful^1^. Patients were arguably identified sooner, and therefore isolated more rapidly. Alternatively, infected individuals passed away before allowing the virus to spread within communities.

Our present working hypothesis is as follows: the P323L RdRp mutation decreases the production of viral RNA and hence the severity of the disease and symptoms. Such a viral variant would be more difficult to detect but would also have a disadvantage with respect to transmission. The D614G S-protein mutation would allow the virus to be transmitted even by an asymptomatic individual because of its ability to infect cells more efficiently, which would allow the transmission of the disease even if only few virions are present.

What do we know about the underlying molecular mechanisms? Regarding the S-protein, the analysis of publicly available simulations reveals an interesting interaction network around the D614 residue. The main interactions change between the two molecular dynamics datasets, observing a cooperative effect among protomers following the symmetry of the oligomer. We believe that the mutation of D614 will disrupt the interactions that couple the AC, CB and BA interfaces. In particular, none of the salt bridges related to D614 could be formed, shifting the conformational population to a more open-like situation (Figure 1a-b). Therefore, the D614G mutation observed more prevalently in specific regions might help to reduce the energetic cost of the conformational transition needed to attach to the cell receptors. In the case of the RdRp, the P323L mutation is located far from the RNA binding domain, which lies within the finger-palm-thumb domains and, therefore, any direct effect of the mutation to the ability of the mutated RdRp to bind RNA should be excluded. However, the mutation could exert its effect allosterically. Since allosteric effects are impossible to be inferred by static crystal structures, simulation techniques can be employed to further elucidate the effect of the mutation on the dynamics of the interface domain and the NiNAR, fingers, and palm domains that are in direct contact with the domain where the mutation is found. Moreover, experiments on the stability of the P323L variant are needed, since changes in the stability of the mutated nsp12 could alter its cellular concentration, which would subsequently affect the replication rate of the virus within the host cell.

Taken together, our study demonstrates that the mutation D614G in S-protein has high infectivity in cellular systems and provides a structural explanation of this effect through molecular modelling. Our data, however, are not compatible with the concept that the D614G S-protein mutation by itself is sufficient to explain the epidemiological success of the presently predominant SARS-CoV-2 strain. The coevolution of the S-protein mutation with the P323L mutation of the RdRp is striking and appears to be crucial for the success of the viral strain. We provide a working hypothesis where the polymerase mutation attenuates the symptoms and severity of the infection, while the spike mutation improves the ability of the D614G spike variant to infect cells.

## Methods

### Cell lines and culture

Cells were maintained in Dulbecco modified Eagle medium (DMEM) supplemented with 10% (v/v) fetal bovine serum (FBS), 100 μg/ml of penicillin and streptomycin, 2 mM l-glutamine, 1 mM sodium pyruvate and 1% non-essential amino acids. Cultures were maintained at 37°C in a 5% CO2 atmosphere. Human cell lines used in the study were HEK 293T/17 (human kidney, ATC C#CRL-11268), A549 cells and HeLa cells

### Luciferase reporter assay

Cells were transduced with SARS-CoV-2 S-protein lentiviral vectors encoding GFP and Gaussia luciferase (Gluc). Four to five days later, GLuc activity was measured by adding 5 uL of the cell supernatant sample to 50 uL of room temperature assay buffer (100 mM NaCl, 35 mM EDTA, 0.1% Tween 20, 300 mM sodium ascorbate, 4 μM coelenterazine in 1X phosphate-buffered saline [PBS]) and immediately measuring luminescence in a luminometer (Glomax; Promega).

### Generation of cell lines expressing ACE2 and TMPRSS2

HeLa and A549 cells were plated at a density of 2E05 cells per well in a 12 well plate. The day after cells were co-transduced either with ACE2/puromycin and/or TMPRSS2/blasticidin lentiviruses. Four day after transduction, cells were selected with blasticidin amd puromycin at 5 μg/ml. Cells were maintained as polyclonal population.

### Plasmids

ACE2 and TMPRSS2 cDNA ORFs were purchased from GenSript and were cloned into pCDHCMV-MCS-EF1α-Puro and pCDH-CMV-MCS-EF1α-Blast lentivectors respectively using standard cloning methods. pCG1_SCoV-2 plasmid encoding SARS-CoV-2 S-protein was provided by Prof. Dr. Stefan Pöhlmann (Deutsches Primatenzentrum, Leibniz-Institute for Primate research, Göttingen, Germany). G614 S-protein was cloned using site-directed mutagenesis with the following primers: 5’-GCGTGAACTGTACCGAAGTG-3’ and 5’-CCTGGTACAGCACTGCC-3’. C-terminal 19 amino acids truncated version of the SARS-CoV-2 S-protein was generated by PCR amplification using primers 5’-AGCGAATTCGGATCCGCC-3’ and 5’-ACAGTCGACTCTAGATTAGCAGCAGCTGCCACAG-3’ followed by cloning into pCG1 plasmid.

### Lentivirus production

For recombinant-lentivirus production, plasmids were transfected in 293T cells using the calcium phosphate method. Briefly, 4.5×10^6^ cells were plated in a 10-cm dish and transfected 16 h later with 15 μg either ACE2, TMPRSS2, or empty-expressing lentiviral vectors, 10 μg of packaging plasmid (psPAX2, gift from Didier Trono [Addgene plasmid 12260]), and 5 μg of envelope (pMD2G, gift from Didier Trono [Addgene plasmid 12259]). The medium was changed 8 h post-transfection. After 48 h, the viral supernatants were collected and filtered using 45 µm PVDF filters and stored at –80°C.

### SARS-CoV-2 S-protein pseudotyped lentiviral production

Spike-pseudotyped lentivirus were produced by co-transfection 293T cells with psPAX2, pCDH-CMV-Gluc-EF1α- GFP, and plasmids encoding either SARS-CoV-2 S-protein D614 full length (FL), S-protein G614 full length (FL), S-protein D614 DeltaCter (DL), S-protein G614 DeltaCter (DL) or empty vector by using the calcium phosphate method as described above.

### SARS-CoV-2 S-protein pseudotyped lentiviral infectivity assays

HeLa and A549 cells were seeded into 96-well plates at a density of 1000 cells per well and inoculated with 50 μL media containing S-protein pseudovirions. After two-day incubation, cells were washed with PBS and the media was refreshed. Two days later, supernatants were collected to measure Gaussia luciferase activity. The cells were trypsynized and resuspended in PBS and were analysed by flow cytometry using Attune NxT Flow Cytometer (Thermo Fisher Scientific). Data were analysed using FlowJo 11 (FlowJo, LLC, Ashland, OR)

### Real-time quantitative RT-PCR

Total RNA was extracted from A549 and HeLa cell lines using RNeasy Micro Kit (Qiagen). Complementary DNA was synthesized from total RNA using PrimeScript™ RT Reagent kit (Takara). The real-time PCR measurement of cDNAs was performed using PowerUp SYBR™ Green Master Mix (Applied Biosystems) and normalized to the expression of GAPDH as control housekeeping gene. The primers were: Furin forward: ggaacatgacagctgcaact; Furin reverse: tcgtcacgatctgcttctca; Cathepsin L forward: tagaggcacagtggaccaag; Cathepsin L reverse: atggccattgtgaagctgtg; ACE2 forward: cattggagcaagtgttggatctt; ACE2 reverse: gagctaatgcatgccattctca; TMPRSS2 forward: cacggactggatttatcgacaa; TMPRSS2 reverse: cgtcaaggacgaagaccatgt; GAPDH forward: gcacaagaggaagagagagacc; GAPDH reverse: aggggagattcagtgtggtg.

### Epidemiological data

Metadata and 36,105 sequences specified as ‘complete’ and ‘high coverage’ were retrieved from GISAID (http://www.gisaid.org/)^23^ as of 30 June 2020. Only sequences that had metadata information were filtered further to contain human as host, a full date including month of the day and that were collected before 22 June 2020, leaving a total of 34,841 sequences. The latter, together with the reference sequence (NC_045512.2), were used to perform multiple sequence alignment (MSA) using MAFFT^4^ (v7.453) with default parameters. We retained only sequences where the first amino acid in S and RdRp protein were identical to the reference proteins, respectively. Next we filtered out sequences that contained non-nucleotide bases e.g. ‘N’ and whose sequence length was different from the respective reference gene. We kept only sequences that corresponded to these criteria for both S protein and RdRp, leaving a total of 27,084 sequences. Among these there were two variants for both S-protein and RdRp protein representing the vast majority of sequences (Supplementary Figure 4). Due to the very low number of other sequence variants (N614 S-protein and F323, H323, S323 in the RdRp protein), we removed sequences containing other variants, leaving us with 27 075 sequenced genomes for further analysis.

Unless stated otherwise only countries that had sequenced a minimum of 20 genomes were used in the analysis. However, for Worldwide analysis, all countries were included. All plots were done using ggplot2 in R. The correlation plots were done using ggscatter function from the ‘ggpubr’ library. The statistical interference between the two dominant variants was performed using pairwise ‘wilcox.test’ function.

### S-protein modelling

The simulations of S-protein used the Amber ff99SB-ILDN force field for proteins^27^, the CHARMM TIP3P model^28^ for water, and the generalised Amber force field^29,30^ for glycosylated asparagine. The system was neutralized and salted with 0.15 M concentration of NaCl. The interval between frames of the analysed trajectories is 1.2 ns, and the simulations were run at 310 K in the NPT ensemble.

## Data Availability

The epidemiological data in this manuscript was retrieved from the public domain (GISAID).

## Acknowledgements

F.L.G. acknowledges funding from Engineering and Physical Sciences Research Council (Grants EP/P022138/1, EP/P011306/1) and European Commission H2020 Human Brain Project CDP 6. HEC-BioSim (EP/R029407/1), PRACE (COVID-19 Fast Track Call), the Swiss National Supercomputing Centre (Project S847 and Project 86), and the HLRS Supercomputing Center (HAWK, PrCov45 and PrCov36) are acknowledged for their generous allocation of supercomputer time. We also are grateful to GISAID and its team for creating a resource for SARS-CoV-2 and to the many participants who kindly provide the data.

## Supplementary Figures and Table

Supplementary Figure 1. Heat maps representing the pairwise root-mean-squaredeviation (RMSD) matrices for RDB and the NTD domains of each the protomers of the wild-type S-protein in the open (magma palette) and closed conformation (virdis palette). Pairwise RMSD matrices were calculated using the python package MDAnalysis^31-34^.

Supplementary Figure 2. Heat maps representing the pairwise root-mean-square-deviation (RMSD) matrices for RDB and the NTD domains of each the protomers of the D614G S-protein in the open conformation (magma palette).

Matrix calculated from a 600ns molecular dynamics simulation initiated in the ‘up’ state (PDBid 6VXX) replacing D614 for glycine in the three protomers of the oligomer. For consistency the simulations used the Amber ff99SB-ILDN force field^27^ for proteins, the TIP3P model^28^ for water, and the Amber Glycam force field^29,30^ for the glycosylated parts of the system. Pairwise RMSD matrices were calculated using the python package MDAnalysis^31-34^.

Supplementary Figure 3. Statistical comparison of the patients’ statuses between D614/P323 and G614/L323. For 1 845 sequences, patient status at the time of sampling was available (http://www.gisaid.org/^23^; only 1 status per patient). **(c) solid bars:** number of patients with status reported as “asymptomatic”, “deceased”, “home”, “hospitalised”, “live”, “recovered”, “released“, “symptomatic”, or “unknown”; **transparent bars:** total number of sequences in countries having reported the respective patient status. Only statuses with at least 50 occurrences are shown. The statistical significance was calculated using Fisher’s Exact Test for Count Data in R. The distribution of patients’ statuses in different countries is shown in Supplementary Figure 6.

Supplementary Figure 4. Worldwide counts of specific amino acid variants in S-protein and RdRp.

Supplementary Figure 5. Weekly number of mutation variants of both double and single mutations in different countries. Only countries with at least 20 sequences are shown.

Supplementary Figure 6. Distribution of patients’ statuses in different countries. Only statuses that occurs at least 50 times are shown.

**Supplementary Table 1. GISAID^23^ acknowledgment table**.

## References

1 Petersen, E. et al. Comparing SARS-CoV-2 with SARS-CoV and influenza pandemics. The Lancet. Infectious diseases, doi:10.1016/s1473-3099(20)30484-9 (2020).

2 Smith, E. C., Blanc, H., Surdel, M. C., Vignuzzi, M. & Denison, M. R. Coronaviruses lacking exoribonuclease activity are susceptible to lethal mutagenesis: evidence for proofreading and potential therapeutics. PLoS pathogens 9, e1003565, doi:10.1371/journal.ppat.1003565 (2013).

3 Ferron, F. et al. Structural and molecular basis of mismatch correction and ribavirin excision from coronavirus RNA. Proc Natl Acad Sci U S A 115, E162–E171, doi:10.1073/pnas.1718806115 (2018).

4 Denison, M. R., Graham, R. L., Donaldson, E. F., Eckerle, L. D. & Baric, R. S. Coronaviruses: an RNA proofreading machine regulates replication fidelity and diversity. RNA Biol 8, 270–279, doi:10.4161/rna.8.2.15013 (2011).

5 Yan, R. et al. Structural basis for the recognition of SARS-CoV-2 by full-length human ACE2. Science (New York, N.Y.) 367, 1444–1448, doi:10.1126/science.abb2762 (2020).

6 Wrapp, D. et al. Cryo-EM structure of the 2019-nCoV spike in the prefusion conformation. Science (New York, N.Y.) 367, 1260–1263, doi:10.1126/science.abb2507 (2020).

7 Jaimes, J. A., Millet, J. K. & Whittaker, G. R. Proteolytic Cleavage of the SARS-CoV-2 Spike Protein and the Role of the Novel S1/S2 Site. iScience 23, 101212, doi:10.1016/j.isci.2020.101212 (2020).

8 Korber, B. et al. Spike mutation pipeline reveals the emergence of a more transmissible form of SARS-CoV-2. bioRxiv, 2020.2004.2029.069054, doi:10.1101/2020.04.29.069054 (2020).

9 Becerra-Flores, M. & Cardozo, T. SARS-CoV-2 viral spike G614 mutation exhibits higher case fatality rate. International journal of clinical practice, doi:10.1111/ijcp.13525 (2020).

10 Li, Q. et al. The Impact of Mutations in SARS-CoV-2 Spike on Viral Infectivity and Antigenicity. Cell, doi:10.1016/j.cell.2020.07.012 (2020).

11 Ogawa, J. et al. The D614G mutation in the SARS-CoV2 Spike protein increases infectivity in an ACE2 receptor dependent manner. bioRxiv, doi:10.1101/2020.07.21.214932 (2020).

12 Goodfellow, Goodfellow, I. & Taube, S. I. Calicivirus Replication and Reverse Genetics Viral Gastroenteritis: Molecular Epidemiology and Pathogenesis. 355–378 (2016).

13 Yin, W. et al. Structural basis for inhibition of the RNA-dependent RNA polymerase from SARS-CoV-2 by remdesivir. Science (New York, N.Y.), doi:10.1126/science.abc1560 (2020).

14 Gao, Y. et al. Structure of the RNA-dependent RNA polymerase from COVID-19 virus. Science (New York, N.Y.) 368, 779–782, doi:10.1126/science.abb7498 (2020).

15 Pachetti, M. et al. Emerging SARS-CoV-2 mutation hot spots include a novel RNA-dependent-RNA polymerase variant. Journal of translational medicine 18, 179, doi:10.1186/s12967-020-02344-6 (2020).

16 Hoffmann, M. et al. SARS-CoV-2 Cell Entry Depends on ACE2 and TMPRSS2 and Is Blocked by a Clinically Proven Protease Inhibitor. Cell 181, 271–280.e278, doi:10.1016/j.cell.2020.02.052 (2020).

17 Fukushi, S. et al. Vesicular stomatitis virus pseudotyped with severe acute respiratory syndrome coronavirus spike protein. The Journal of general virology 86, 2269–2274, doi:10.1099/vir.0.80955-0 (2005).

18 Research, D. E. S. Molecular Dynamics Simulations Related to SARS-CoV-2. D. E. Shaw Research Technical Data (http://www.deshawresearch.com/resources_sarscov2.html) (2020).

19 Shaw, D. E. et al. in SC ‘14: Proceedings of the International Conference for High Performance Computing, Networking, Storage and Analysis. 41–53.

20 Katoh, K. & Standley, D. M. MAFFT multiple sequence alignment software version 7: improvements in performance and usability. Molecular biology and evolution 30, 772–780, doi:10.1093/molbev/mst010 (2013).

21 Watanabe, Y., Allen, J. D., Wrapp, D., McLellan, J. S. & Crispin, M. Site-specific glycan analysis of the SARS-CoV-2 spike. Science (New York, N.Y.), doi:10.1126/science.abb9983 (2020).

22 Weissman, D. et al. D614G Spike Mutation Increases SARS CoV-2 Susceptibility to Neutralization. (2020).

23 Elbe, S. & Buckland-Merrett, G. Data, disease and diplomacy: GISAID’s innovative contribution to global health. Glob Chall 1, 33–46, doi:10.1002/gch2.1018 (2017).

24 Dasgupta, B., Chakrabarti, P. & Basu, G. Enhanced stability of cis Pro-Pro peptide bond in Pro-Pro-Phe sequence motif. FEBS letters 581, 4529–4532, doi:10.1016/j.febslet.2007.08.039 (2007).

25 Subissi, L. et al. One severe acute respiratory syndrome coronavirus protein complex integrates processive RNA polymerase and exonuclease activities. Proceedings of the National Academy of Sciences of the United States of America 111, E3900–3909, doi:10.1073/pnas.1323705111 (2014).

26 Holmes, E. C. What does virus evolution tell us about virus origins? Journal of virology 85, 5247–5251, doi:10.1128/jvi.02203-10 (2011).

27 Lindorff-Larsen, K. et al. Improved side-chain torsion potentials for the Amber ff99SB protein force field. Proteins 78, 1950–1958, doi:10.1002/prot.22711 (2010).

28 Jorgensen, W., Chandrasekhar, J., Madura, J., Impey, R. & Klein, M. Comparison of Simple Potential Functions for Simulating Liquid Water. J. Chem. Phys. 79, 926–935, doi:10.1063/1.445869 (1983).

29 Kirschner, K. N. et al. GLYCAM06: a generalizable biomolecular force field. Carbohydrates. J Comput Chem 29, 622–655, doi:10.1002/jcc.20820 (2008).

30 Website builders, e.g., Carbohydrate Builder, Glycoprotein Builder, etc. Woods Group. GLYCAM Web. Complex Carbohydrate Research Center, University of Georgia, Athens, GA. (http://glycam.org) (2005–2020).

31 Beckstein, O., Denning, E. J., Perilla, J. R. & Woolf, T. B. Zipping and unzipping of adenylate kinase: atomistic insights into the ensemble of open<-->closed transitions. J Mol Biol 394, 160–176, doi:10.1016/j.jmb.2009.09.009 (2009).

32 Gowers, R. et al. MDAnalysis: A Python Package for the Rapid Analysis of Molecular Dynamics Simulations. (2016).

33 Michaud-Agrawal, N., Denning, E. J., Woolf, T. B. & Beckstein, O. MDAnalysis: a toolkit for the analysis of molecular dynamics simulations. J Comput Chem 32, 2319–2327, doi:10.1002/jcc.21787 (2011).

34 Theobald, D. Rapid calculation of RMSDs using a quaternion-based characteristic polynomial. Acta crystallographica. Section A, Foundations of crystallography 61, 478–480, doi:10.1107/S0108767305015266 (2005).

